# Epidemiological analysis of all vena cava filter placement over 9 years in Brazil: trends and mortality rates in a population of over 200 million

**DOI:** 10.1101/2025.07.07.25331036

**Authors:** Clara Sanches Bueno, Hedra Marques Santos, Júlia Freire Castanheira, Bruno Jeronimo Ponte, Felipe Soares Oliveira Portela, Marcelo Fiorelli Alexandrino da Silva, Marcelo Passos Teivelis, Alexandre Fioranelli, Nelson Wolosker

## Abstract

**Background:** Venous thromboembolism (VTE) is a major global health concern, with inferior vena cava filters (VCFs) serving as an alternative to anticoagulation in high-risk patients. Brazil’s VCF trends remain understudied, particularly in its dual public-private healthcare system. This database study allows a broad overview of the Brazilian healthcare landscape.

**Methods:** This retrospective nationwide analysis used Brazilian public (SUS/TabNet) and private (ANS/D-TISS) healthcare databases (2015–2023) to assess VCF placement rates, regional disparities and in-hospital mortality. Data included 21,630 procedures, analyzed via generalized linear models (Gamma/Poisson distributions).

**Results:** The private system performed 57% of VCFs (12,323) despite covering only 25% of the population, with a 4.25-fold higher procedure rate per capita than the public system (29.45 vs. 6.42 per million). Women accounted for 57.8% of recipients. The Southeast region dominated (70% private, 40.5% public), while the North and Midwest had the lowest rates. Mortality rates were comparable (public 6.7% vs. private 7.4%, p = 0.066), peaking during COVID-19. Procedure volumes rose 140% over 9 years, contrasting with declining U.S. trends post-FDA restrictions.

Conclusions:

Brazil’s VCF use reflects systemic inequities, with private-system overrepresentation and regional gaps. This study enables unprecedented insights into large-scale VCF implementation across diverse healthcare subsystems.

## INTRODUCTION

Venous thromboembolism (VTE), which includes deep vein thrombosis (DVT) and pulmonary embolism (PE)^1^, represents a significant global health burden, with an estimated 5% lifetime prevalence in adult populations.^2^ The rising incidence of VTE reflects demographic and epidemiological transitions, including aging populations, increasing prevalence of obesity, sedentary lifestyles, and enhanced diagnostic sensitivity through advanced imaging techniques and biomarker assays.^3–4^ Anticoagulation therapy remains the gold standard of VTE treatment. However, vena cava filters (VCF) are an important alternative for specific scenarios, such as when a patient has acute VTE and an absolute contraindication to anticoagulation or when recurrent PE occurs despite adequate anticoagulant therapy.^5–7^

Although the placement of VCFs is a minimally invasive, percutaneous procedure, it is not risk-free. Rare but potentially serious complications can occur, such as filter migration, venous wall penetration, and adjacent organ involvement (e.g., duodenal perforation).^8^ Historically, there has been a trend in the United States toward the overuse of VCFs, with studies indicating that over 50% of procedures were for questionable indications in the early 2000s.^9^ In contrast, studies from the same period in Brazil did not show this trend; VCF placement there was more selective and based on well-established criteria.^10^

Brazil’s universal healthcare system, known as Sistema Único de Saúde (Public), is a publicly funded program designed to provide full-spectrum medical care to all citizens.^11^ Although the constitution guarantees free healthcare access to all citizens, around 25% of the population opts for additional private health services (Private) , often funded through employer-sponsored plans. As a result, approximately 75% of Brazilians rely exclusively on the public healthcare system.^12^

The Public system maintains a national anonymized database that tracks surgical procedures, including all VCF implantations performed in public hospitals. Recently, data on procedures performed within the Private has become publicly available, covering interventions from 2015 to 2023. This new information allows for comparative studies between public and private healthcare systems and provides a broad overview of the Brazilian healthcare landscape. The analysis does not include only out-of-pocket payment data, which represent a minority of expenditures (less than 5%).^13^ Consequently, it enables a thorough assessment of the utilization of VCF in Brazil.

Notably, no population-level study on raw data (as opposed to estimation-based) has yet evaluated VCF placement across an entire nation. Globally, the most extensive analysis of VCF use comes from the U.S. Nationwide Inpatient Sample (NIS), which provides estimates based on a stratified 20% sample of discharges from hospitals participating in the Healthcare Cost and Utilization Project (HCUP).^14,15^ In Brazil, while some studies have examined VCF placement using SUS databases, none have encompassed both Public and Private datasets, leaving a gap in understanding the full scope of procedural trends, disparities, and outcomes.^10,16^

Our study represents a significant advancement by offering the first complete national assessment of VCF utilization patterns, covering all 26 states of this continental-scale nation. Our analysis uniquely incorporates data from both the Public and Private. This approach enables unprecedented insights into large-scale VCF implementation across diverse healthcare subsystems.

**Objectives:** To study all cases requiring VCF placement in Brazil - within the Public and Private - from 2015 to 2023 using a big data system. We intended to analyze variations in the number of procedures over time, trends by geographic region, patient characteristics, and in-hospital mortality rates.

## MATERIALS AND METHODS

This retrospective population-based study analyzed publicly available data from the TabNet platform, of the Department of Informatics of the Unified Health System (DATASUS) and the Private (D-TISS). TabNet provides open-access data on procedures conducted within Public system, which accredited hospitals and outpatient clinics are required to report. Accreditation and data reporting are mandatory for government reimbursement of these procedures. Similarly, the Private healthcare system platform (D-TISS), managed by the National Supplementary Health Agency (ANS), provides open-access procedure data for the Private heath system, excluding out-of-pocket payments.

This research was deemed exempt by the institutional ethics committee and was institutionally registered under the number 10337 (Sistema Gerenciador de Projetos, SGPP). All data provided by DATASUS - TabNet and Private D-TISS are anonymous and publicly available. For this reason, the Institutional Review Council (Conselho de Revisão Institucional, IRB) waived the requirement for informed consent forms.

Data on VCF placement was collected from both platforms (TabNet and D-TISS), covering a 9-year period from 2015 to 2023, excluding 2024 due to incomplete DATASUS data for the period. The collected data included geographic region, the number of procedures performed, in-hospital mortality rates, and financial investments made by both Public and Private. Statistics on in-hospital deaths were selected from the hospital mortality sections of TabNet and D-TISS. The data was grouped by Brazilian geographic region and distributed over the years.

For data extraction within Public, we used a specific code assigned to this procedure established by the Public Procedure, Medicine, and OPM Management System (SIGTAP): 04.06.04.014-1. Data collection for the Private was obtained from the D-TISS Data Panel, published by ANS, and procedures were analyzed according to the Private coding system, which includes Vena cava filter (VCF) placement (code 30907080) and VCF implantation for pulmonary embolism prevention (code 40813240).

The steps for data collection, platform field selection, and table adjustment were performed using the selenium-webdriver packages (v. 3.1.8, Selenium HQ, several collaborators worldwide) and pandas (v. 2.7.13, Lambda Foundry, Inc. e PyData Development Team, New York, USA). The Mozilla Firefox browser (v. 59.0.2, Mountain – California – USA) and geckodriver webdriver (v 0.18.0, Mozilla Corporation, Bournemouth, England) were used. Following collection and treatment, all data were organized and grouped in a spreadsheet using Microsoft Office Excel 2016® (v. 16.0.4456.1003, Redmond – Washington – USA) software. These computer programs allowed automated content access (web scraping), the automated navigation codes were programmed in Python language (v. 2.7.13, Beaverton – Oregon – USA) using the Windows 10 Single Language operating system.

The examined variables included the total number of procedures, patient demographics (gender and age distribution), regional disparities in service provision, and associated mortality rates. For statistical analysis, generalized linear models with a Gamma distribution^17^ were performed to assess the relationship between the rate of procedures and average values with year, source of procedures (public or private), and region. For the death rate, a generalized linear model with a Poisson distribution was used, with the number of procedures as an offset. The results were presented as rate ratios, 95% confidence intervals, and p-values. The analyses were performed using R, version 4.1.1 (R Core Team. R: A Language and Environment for Statistical Computing. 2024. R version 4.4.1).

## RESULTS

Between 2015 and 2023, a total of 21,630 CVF procedures were performed in Brazil, distributed between the Public and the Private. During this period, the Private system accounted for 57% (12,323) of all procedures, while the Public system accounted for 43% (9,307). Notably, despite Private covering only 25% of the population, it performed a significantly higher proportion of CVF procedures. (**Table 1**) ^12^

**Table 1.**
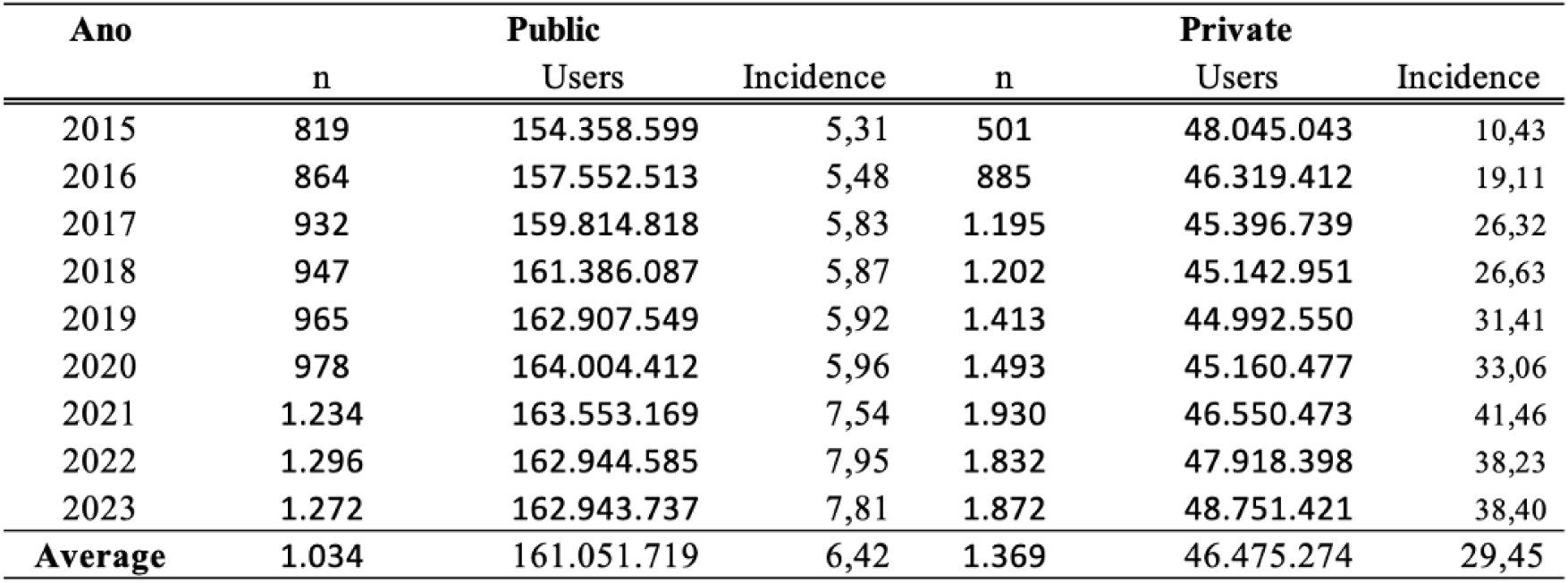
Absolute number of CVF procedures performed and incidence per 1 million beneficiaris in Public and Private health system betwen 2015 and 2023

| Ano | Public |  |  | Private |  |  |
| --- | --- | --- | --- | --- | --- | --- |
|  | n | Users | Incidence | n | Users | Incidence |
| 2015 | 819 | 154.358.599 | 5,31 | 501 | 48.045.043 | 10,43 |
| 2016 | 864 | 157.552.513 | 5,48 | 885 | 46.319.412 | 19,11 |
| 2017 | 932 | 159.814.818 | 5,83 | 1.195 | 45.396.739 | 26,32 |
| 2018 | 947 | 161.386.087 | 5,87 | 1.202 | 45.142.951 | 26,63 |
| 2019 | 965 | 162.907.549 | 5,92 | 1.413 | 44.992.550 | 31,41 |
| 2020 | 978 | 164.004.412 | 5,96 | 1.493 | 45.160.477 | 33,06 |
| 2021 | 1.234 | 163.553.169 | 7,54 | 1.930 | 46.550.473 | 41,46 |
| 2022 | 1.296 | 162.944.585 | 7,95 | 1.832 | 47.918.398 | 38,23 |
| 2023 | 1.272 | 162.943.737 | 7,81 | 1.872 | 48.751.421 | 38,40 |
| **Average** | **1.034** | **161.051.719** | **6,42** | **1.369** | **46.475.274** | **29,45** |

Analyzing the incidence of CVF procedures per 1 million beneficiaries (**Table 1**), the private system exhibited a consistently higher incidence rate (average of 29.45 per 1 million beneficiaries) compared to the public system (average of 6.42). While the absolute number of procedures in the public system showed moderate growth (from 819 to 1,272), the private system experienced a sharp increase (from 501 to 1,872), peaking in 2021 (incidence: 41.46). Notably, the public system’s incidence rose during the pandemic (7.45 in 2021).

A predominance of women was observed in CVF procedures across both systems (**Table 2**). In Public, out of 9,307, 59.15% were women (5,506 cases) and 40.84% men (3,801 cases). In Private (12,323), 56.25% were women (6,932 cases) and 42.75% men (5,268 cases). Overall, of 21,507 procedures with reported gender, 57.83% were women (12,438 cases) and 42.17% were men (9,069 cases). Gender was unreported in only 123 cases (0.57%).

**Table 2.** Distribution by gender of CVF placement

| Sexo | Public |  | Private |  | Total |  |
| --- | --- | --- | --- | --- | --- | --- |
|  | n | % | n | % | n | % |
| Men | 3801 | 40,84 | 5268 | 42,75 | 9069 | 41,93 |
| Women | 5506 | 59,15 | 6932 | 56,25 | 12438 | 57,5 |
| Not informed |  |  | 123 | 1 | 123 | 0,57 |

Figure 1 presents the age distribution of procedures in Public and Private. In Public, the largest share (23.98%) fell in the 60-69-year-old group. By contrast, in Private, the over-80-year-old cohort accounted for the highest proportion (2,718 cases; 22.06%), versus 751 Public cases (8.07%). Additionally, 122 Private procedures (0.99%) lacked age data.

**Figure 1.**
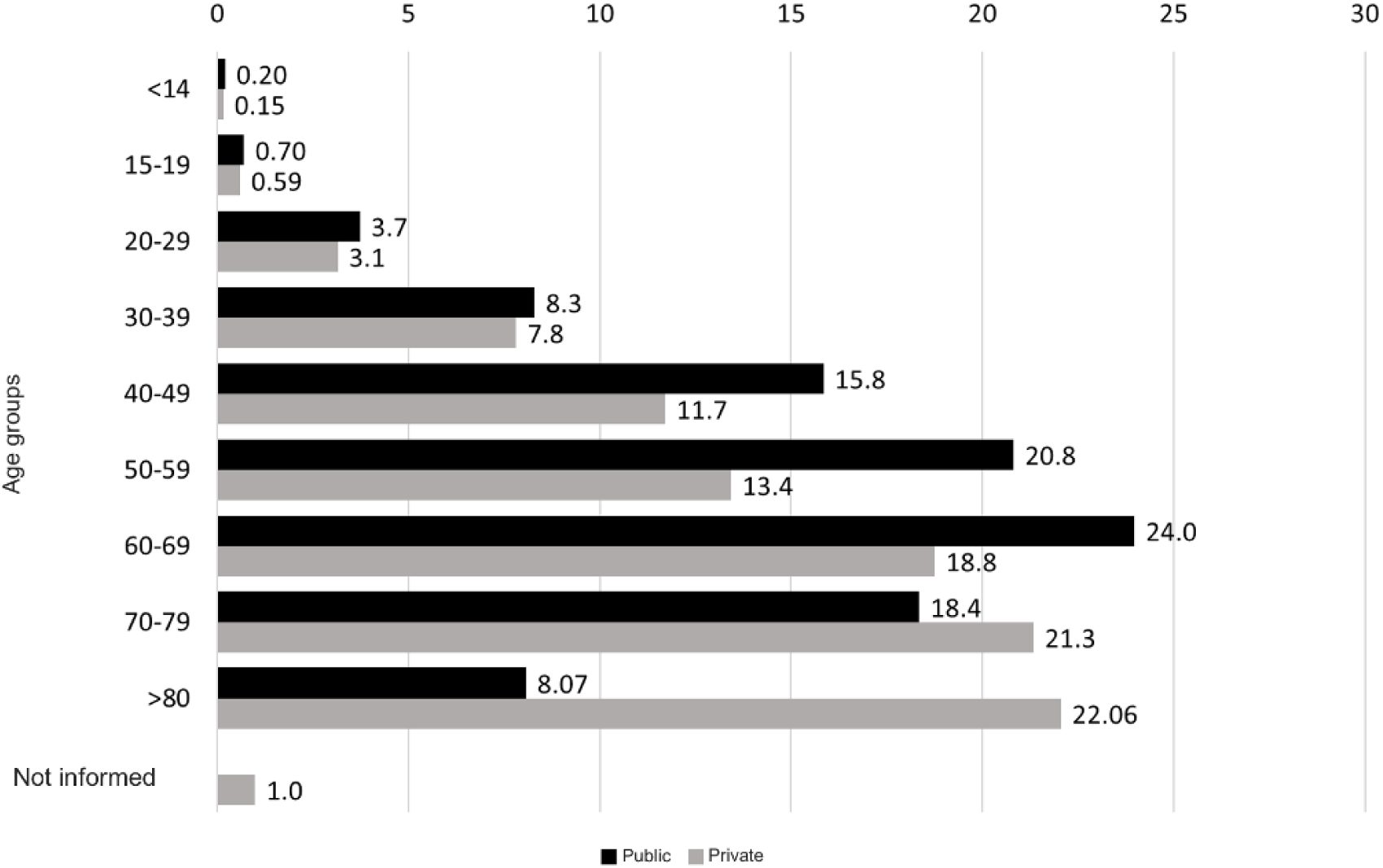
Distribution by age group of procedures carried out in Public and Private healthcare systems

**Table 3** presents the procedures performed by region. In Public, the Southeast led with 3,771 procedures (40.518%), followed by the Northeast with 3,074 (33.029%). In Private, the Southeast region again dominated with 8,620 procedures (69.950%), then the Northeast with 1,942 (15.759%), the South with 1,033 (8.383%).

**Table 3.** Procedures by region of the country

| Region | Public |  | Private |  | Total |  |
| --- | --- | --- | --- | --- | --- | --- |
|  | n | % | n | % | n | % |
| North | 128 | 1,38 | 163 | 1,32 | 291 | 1,345 |
| Northeast | 3.074 | 33,03 | 1.942 | 15,76 | 5.016 | 23,190 |
| Midwest | 332 | 3,57 | 565 | 4,58 | 897 | 4,147 |
| Southeast | 3.771 | 40,52 | 8.620 | 69,95 | 12.391 | 57,286 |
| South | 2.002 | 21,51 | 1.033 | 8,38 | 3.035 | 14,031 |
| Total | 9.307 | 100,00 | 12.323 | 100,00 | 21.630 | 100,000 |

Both systems show a predominance of procedures in the Southeast, substantial contributions from the Northeast and South, and minimal activity in the Midwest and North a pattern consistent over time in the Brazil areas with the lowest demographic density. (Figure 2)

**Figure 2.**
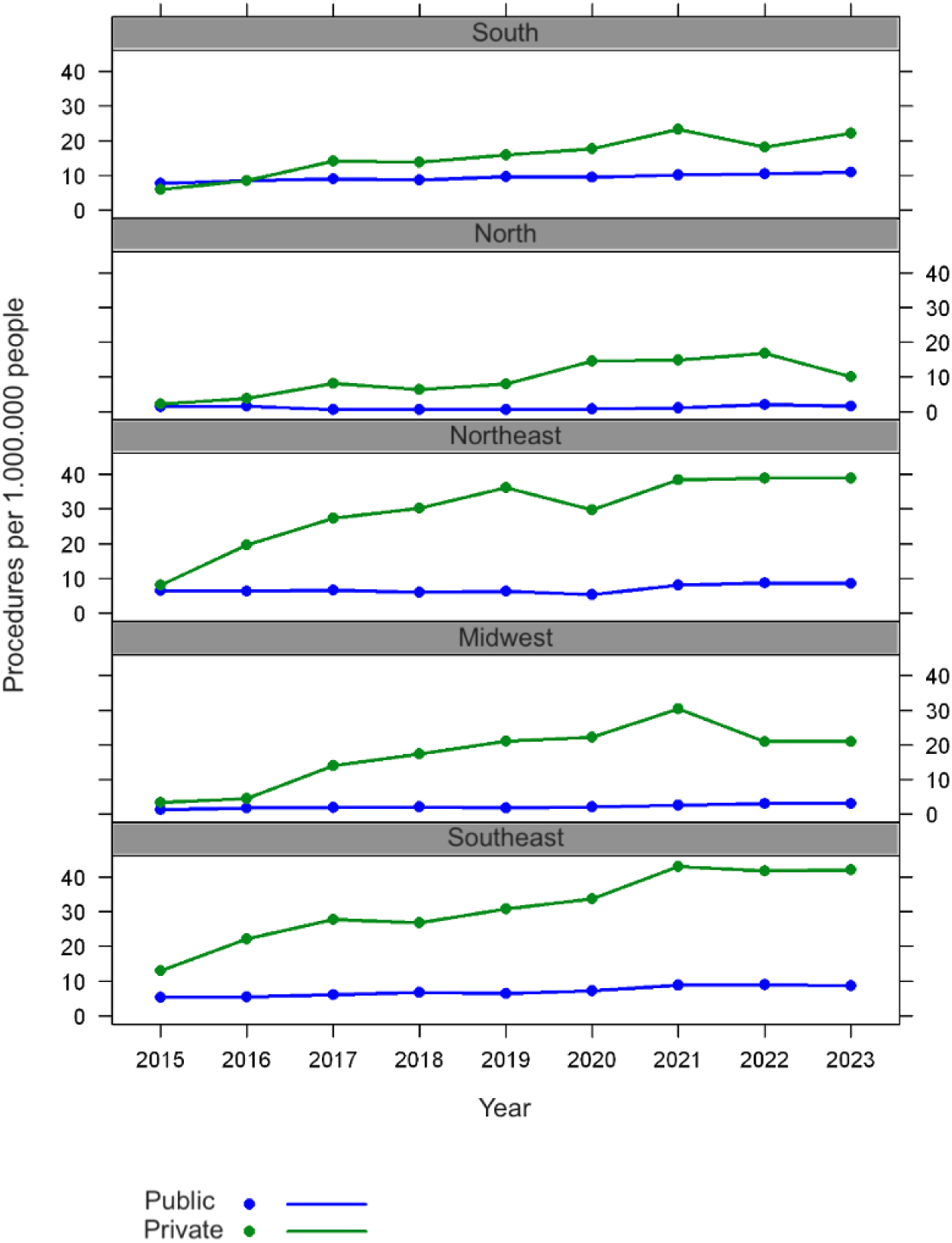
Procedures per 1.000.000 people

The adjusted model for the procedure rate, accounting for the region of origin, shows a significant change over the years (p-value < 0.001), with an estimated annual increase of 11% in the rate. Regarding the source of care, the procedure rate under Private is estimated to be 4.357 times higher than Public (p-value < 0.001) (**Table 4**).

**Table 4.** Procedure rate for 1.000.000 patients

|  | Mean rate (CI 95%) | p-value |
| --- | --- | --- |
| Year | 1.110 (1.074; 1.149) | <0.001 |
| Source |  |  |
| Public | Reference |  |
| Private | 4.357 (3.654; 5.193) | <0.001 |
| Region |  |  |
| Southeast | Referência |  |
| Midwest | 0.424 (0.325; 0.553) | <0.001 |
| Northeast | 0.971 (0.745; 1.267) | 0.831 |
| North | 0.225 (0.172; 0.294) | <0.001 |
| South | 0.931 (0.712; 1.218) | 0.600 |
| Not adjusted by region |  |  |
| Source |  |  |
| Public | Reference |  |
| Private | 3.652 (2.883; 4.626) | <0.001 |

CI: confidence interval

Analyzing the mortality associated with VCF implantation procedures (**Table 5**), overall rates were 7.4% in Private (918 / 12,323 procedures) versus 6.7% in Public (625 / 9,307 procedures). In Public, regional mortality rates ranged from 3.1% in the North to 12.7% in the Midwest; in Private, from 3.7% in the North to 8.1% in the Northeast. The largest Public-Private disparity appeared in the Midwest (12.7% vs. 6.4%). In both the Southeast (7.0% vs 7.4%) and South (6.7% vs. 7.4%).

**Table 5.** Absolute and relative distribution of deaths of patients undergoing surgical treatment per region for CVF placement in the SUS and PHS

| Region | Public |  |  | Private |  |  |
| --- | --- | --- | --- | --- | --- | --- |
|  | Procedures | Deaths | Mortality rate | Procedures | Deaths | Mortality rate |
| North | 128 | 4 | 3,1 | 163 | 6 | 3,7 |
| Northeast | 3.074 | 181 | 5,9 | 1.942 | 158 | 8,1 |
| Midwest | 332 | 42 | 12,7 | 565 | 36 | 6,4 |
| Southeast | 3.771 | 264 | 7 | 8.620 | 642 | 7,4 |
| South | 2.002 | 134 | 6,7 | 1.033 | 76 | 7,4 |
| Total | 9.307 | 625 | 6,72 | 12.323 | 918 | 7,45 |

Private rates were slightly higher than Public. However, the adjusted model for the mortality rate showed no evidence of differences between healthcare sources (public vs. private, p-value = 0.066) and no significant change over the years (p-value = 0.053). A lower mortality rate in the North region compared to the reference region was noted (**Table 6**).

**Table 6.** Mortality rate per 1,000 procedures

|  | Mean rate (CI 95%) | p-value |
| --- | --- | --- |
| Year | 1.020 (1.001; 1.041) | 0.056 |
| Source |  |  |
| Public | Reference |  |
| Private | 1.086 (0.975; 1.209) | 0.134 |
| Region |  |  |
| Southeast | Reference |  |
| Midwest | 1.191 (0.937; 1.490) | 0.139 |
| Northeast | 0.952 (0.836; 1.082) | 0.453 |
| North | 0.474 (0.237; 0.835) | 0.019 |
| South | 0.978 (0.836; 1.139) | 0.779 |
| Not adjusted by region |  |  |
| Source |  |  |
| Public | Reference |  |
| Private | 1.100 (0.994; 1.219) | 0.066 |

CI: confidence interval

## DISCUSSION

This retrospective study utilizes population data from TabNet (DATASUS) and D-TISS (ANS) to assess trends in VCF use in Brazil. Rather than analyzing a sample, it covers a great part of Brazilian population over nine years from 2015 to 2023. It is the first study to analyze the entire population of a continental developing country.

A total of 21,503 VCFs were placed during this period: 9,307 in Public and 12,196 in Private. While Public averaged 6.42 procedures per 1 million beneficiaries, Private - serving only 25% of the population - had a fivefold higher rate (29.45 per 1 million). This contrasts with U.S. data for Medicare beneficiaries, which showed a decrease from 1578.0 to 641.0 per million (2010 and 2021)^18^ due to stricter guidelines – yet peak U.S. rates were still 20 times higher than Brazil’s private system.

European data show intermediate usage: in 2012, the “Big Five” nations (France, Germany, Italy, Spain, and the UK) reported 9,070 VCF placements or 28 per million.^19^ These contrasts reflect major differences in clinical practice across healthcare systems.

It is also noteworthy that although Private has significantly higher rates compared to Public in Brazil, it remains lower than those observed in many high-income countries around the world.

The fivefold higher VCF rate in Brazil’s Private compared to Public suggests a strong economic influence. Individuals with health insurance generally access endovascular procedures more easily, as high-resolution imaging and endovascular surgery are more available in private centers.^20^

Placing a VCF requires a team trained in endovascular surgery, more often found in private hospitals or hospitals or private-affiliated hospitals. Within Public sector, high-complexity services are less available, especially in the North and Northeast regions, limiting access to such procedures. ^21^

Private healthcare systems typically operate under reimbursement models that may encourage more procedures. Although no direct evidence confirms financial drivers for VCFs in Private, fee-for-service models likely affect decisions, unlike the Public’s limited budget.^22^ Demographics in the Private population (older, more comorbid, higher socioeconomic status) might slightly influence VTE risk, but these factors alone do not explain the large gap in VCF usage. ^23,24^

Recent trends in VCF placement show a clear contrast between the United States and Brazil. In the U.S., VCF use significantly declined after FDA alerts in 2010 and 2014, and the implementation of stricter guidelines aimed at correcting overuse and emphasizing filter retrieval – dropping among Medicare beneficiaries from 1578 to 641 insertions per 1 million between 2010 and 2021. ^18^Conversely, Brazilian data from 2015 to 2023 indicate a general upward trend in both the absolute number of procedures (Private + Public, from 1,320 in 2015 to 3,144 in 2023) and in rates per million inhabitants, particularly in the private healthcare sector, which rose from 10.18 to 38.34 per million, and also in Public, which increased from 5.31 to 7.81 per million, with a more pronounced increase from 2021 onwards.

Several hypotheses can explain these trends. Firstly, Brazil began with a considerably lower VCF usage baseline than the U.S., and its 140 % increase over eight years might reflect an expansion of access to appropriate technology as its healthcare system evolves. Secondly, a rising incidence of VTE, possibly worsened by the COVID-19 pandemic and its known VTE risk, may also have contributed to the post-2021 surge.^25^

A clear finding is that VCF procedures were more frequent in women, who accounted for 57.50% (n = 12,438) of all VCF placements, versus 41.93% (n = 9,069) in men. This trend was observed in both healthcare settings: 59.16% in Public and 56.25% in Private. These consistent results suggest that the conditions requiring VCFs are more prevalent in women, independent of healthcare coverage.^10,26^

This finding may reflect an increased risk of PTE in women exposed to hormonal contraceptives, pregnancy, postpartum, or hormone replacement therapies.^10,18^

VCF placement is primarily age-associated, with approximately 74% of procedures in patients aged 50 or older. This trend is attributed to the higher risk of PTE due to chronic conditions like cancer and kidney disease.^27–29^ Additionally, older individuals often experience mobility limitations, especially following major surgeries, and frequently use medications that affect coagulation, such as hormone replacement therapies and oncological treatments. ^5^

Age group analysis reveals differences between Public and Private. In Public, procedures concentrate in ages 60-69 (23.98%), and 50-59 (20.82%). In Private, the distribution skews older, with 80+ (22.06%) and 70-79 (21.34%) as the leading groups. The predominance of very advanced age groups (>70 and particularly >80 years) in the Private may suggest that individuals with higher socioeconomic status, who are more likely to have private health insurance and greater longevity, seek or have facilitated access to specific procedures within the private sector during this life stage. In contrast with Public patients witch frequently have a low-income status and may have reduced longevity.

An analysis of the regional distribution of healthcare procedures in Brazil reveals that the Southeast region accounts for most procedures, with 40.5% in the Public and 70% in the Private. This concentration can be attributed to the region’s higher population density, widespread availability of supplementary health plans, and a larger pool of specialized healthcare professionals.^24^ In contrast, the Midwest and North regions account for less than 5% of Public procedures and 6% of Private procedures, highlighting significant gaps in healthcare access, also reflecting demographic void. These disparities may lead to patients needing to travel interstate for complex cases and could necessitate the placement of VCFs.

The comparative analysis of in-hospital mortality following VCF placement in Brazil (2015-2023) indicates that although the Private performed more procedures and consequently recorded a higher absolute number of deaths, its mortality rate was generally higher that of the Public (7.42% vs. 6.63%). In 8 of 9 years analyzed, Private rates exceeded those of Public (2022 was the exception, and 2017 showed similar rates). Both systems exhibited fluctuations and peaks - especially between 2020 and 2022 – likely influenced by the COVID-19 pandemic. However, these raw figures do not account for confounders such as case severity and complexity, differing eligibility criteria, and patient demographics. For example, Private patients may have access to more complex procedures or be transferred in more critical condition, while Public patients might have multiple comorbidities and face delayed access to care. Consequently, any inference about care quality requires a risk-adjusted analysis, which is currently unfeasible due to unavailable data.

Overall, the in-hospital mortality rate for VCF placement is similar in both systems (6.7% in Public vs. 7.4% in Private). These rates largely reflect patients’ underlying severe conditions (e.g., VTE, cancer, or trauma), whereas mortality directly attributable to the VCF implantation procedure itself is usually below 1%,^8,30^ as shown in systematic reviews and clinical trials such as PREPIC^18^. Direct comparisons across studies are complex due to variations in patient profiles, filter types, indications (therapeutic vs. prophylactic), and the definitions and follow-up periods for mortality. Regional variations observed in Brazil (Public rates ranging from 3.1% to 12.7% and Private rates from 3.7% to 8.1%) also suggest the influence of local factors and specific characteristics of each region’s population.

The mortality rates in Brazil between 2015 and 2023 find parallels in international studies for severe cohorts. In a large observational study in the USA (SAFE-IVC, with Medicare data from 2013-2021) found a 30-day all-cause mortality rate of 14.6% in patients who received VCF; Brazil’s 6.7% and 7.4% rates are lower.^18^

## LIMITATIONS

This study has inherent limitations due to the use of aggregated administrative databases (DATASUS and D-TISS), which are subject to coding errors, reporting inconsistencies, and potential misclassification or underreporting. Data anonymization prevented longitudinal tracking of individual patients, so unique patient counts cannot be distinguished from total VCF procedures, as one patient may have undergone multiple events (implantation, removal, reintervention). Also, the age stratification of all users of private system was not available, so age-specific rates could not be calculated. Furthermore, out-of-pocket procedures, though likely representing a smaller proportion of cases, were not captured in this analysis.

Also, inconsistencies in coding and reporting quality made it impossible to categorize patients by comorbidities or clinical severity, limiting risk-factor analysis.

Despite these limitations, this study offers a novel, comprehensive nationwide overview of VCF utilization in Brazil (2015–2023) across both public and private healthcare systems. By consolidating official data, it provides valuable real-world insights into differences in procedure volumes thereby contributing to a better understanding of clinical practice and financing of this technology in the country.

## CONCLUSIONS

In this analysis based on real-world data from 2015 to 2023, a total of 21,630 VCF implantation procedures were performed in Brazil. This resulted in an average annual rate of 1.16 procedures per 10,000 inhabitants, considering the combined estimated populations covered by the Public and the Private.

The procedure rate per 10,000 inhabitants in the Private was 4.25 times higher than in the Public.

## Data Availability

All data produced in the present work are contained in the manuscript

